# Impact of Obesity and Blood Pressure on Left Ventricular Hypertrophy in Adults with Congenital Heart Disease

**DOI:** 10.1101/2025.08.06.25333174

**Authors:** Matthew Laubham, Aaron T. Walsh, Kan N. Hor, May Ling Mah, Chance Alvarado, Andrew H. Tran

## Abstract

**Background:** The majority of children with congenital heart disease (CHD) survive into adulthood, and acquired cardiovascular risk factors, such as hypertension and obesity, are increasing considerations for adults with congenital heart disease (ACHD). Hypertension and obesity are associated with the development of left ventricle hypertrophy (LVH) in the general population. This study aims to evaluate the impact of obesity and blood pressure on LVH in the ACHD population.

**Methods:** We retrospectively analyzed echocardiograms from subjects with biventricular CHD aged >18 years from 2012-2019. CHD lesion types were grouped according to the original embryologic pre-repaired form. We defined LVH using indexed cutoff values of ≥ 51 g/Ht^2.7^, and >115 g/m^2^ for males and >95 g/m^2^ for females. Patients were grouped by blood pressure (BP) into normotensive (NT, systolic BP (SBP) < 120 mm Hg), Elevated BP (E-BP, 120 ≥ SBP < 130 mm Hg), Stage 1 HTN (HTN-1, 130 ≥ SBP < 140 mm Hg), and Stage 2 HTN (HTN-2, SBP ≥ 140 mm Hg). Obesity was defined as underweight (BMI <18.5), healthy weight (BMI 18.5≤ - <25), overweight (BMI 25≤ - 30), and obese (BMI ≥30).

**Results:** There were 1,152 subjects included. Median age was 24 years (IQR 20, 32), with 50% females, median SBP of 121 mmHg, and median LVM of 36 g/ Ht^2.7^ and 78 g/m^2^. There were 519/1152 (45%) in the NT group, 313/1152 (27%) E-BP, 183/1152 (15.8%) HTN-1, and 137/1152 (12%) HTN-2. When evaluating BMI, a model adjusting for confounders of age, sex, and cardiac diagnosis, demonstrated that a 10-unit increase of BMI was associated with an increase in LVM indexed by BSA of 1.27 g/m^2^ (95% CI: −0.38, 2.93; p = 0.13), and 7.95 g/m^2.7^ (95% CI: 7.13, 8.77; p < 0.001) when indexed to LVH-Ht^2.7^. BMI was strongly associated with increased in LVH-Ht^2.7^, as having knowledge of an individual’s BMI showed a 21% increase in explained variation.

**Conclusions:** Obesity demonstrated the highest level of correlation on the development of LVH in this ACHD population, as it demonstrated a 21% increase in explained variation. This finding highlights the need for early prevention and weight loss interventions in the ACHD population.

## Introduction

Congenital heart disease (CHD) remains the most common birth defect in the United States each year. ^1^ Globally, the prevalence of CHD is estimated to be 9.1 / 1000 births, and accounts for one-third of all congenital anomalies. ^2^ With many improvements in medical, catheter-based and surgical techniques, more patients born with CHD are living past the first decade of life and beyond. In developed countries, more than 90% of patients with CHD survive into adulthood such that adults with CHD (ACHD) now comprise two-thirds of the total CHD population. ^3, 4^

An aging ACHD population faces many of the same well recognized modifiable risks that adults without CHD encounter that impact the development of acquired cardiovascular (CV) disease, including metabolic syndrome, hypertension (HTN), and obesity. ^5–7^ While CV risks increase with age in both adults with and without CHD, ACHD patients may manifest several cardiovascular risk factors at rates higher than the general adult population, including systolic hypertension (HTN) and arterial stiffness. ^6, 8, 9^ Furthermore, hospital admissions for acquired CV disease including coronary disease have increased among adults with CHD over more than a decade. ^10^

It is well described that both hypertension and obesity are important factors associated with the development of adverse left ventricle remodeling and function in the general population.^11–15^ To that end, increases in LVM is also associated with adverse cardiovascular events. ^16–21^

Similar to adults without CHD, hypertension and obesity are considered cardiovascular risks for ACHD patients, and both risks impact the development of LVH. Moreover, lead time impacts may also contribute, as adolescents with CHD have a higher prevalence of HTN and LVH, and children with CHD can be at higher risk for future CV events compared to non-CHD children. ^22, 23^ There is much less known about the prevalence of LVH in a wide variety of ACHD patients, however, as the intersection of the impact HTN and obesity has on LVH has largely been reported in congenital heart disease patients with left sided obstructive lesions. ^24–26^ In ACHD populations, there is currently no defined risk factor profile that contributes LVH. Therefore, the aim of our study was to determine the impact of obesity and HTN on LVH in adults with a wide array of congenital heart disease defects.

## Methods

### Study Population

We performed a retrospective analysis of patients aged 18 years and older with biventricular CHD from 2012-2019 at a single-center quaternary care center with a large ACHD population. The Nationwide Children’s Hospital Institutional Review Board approved the protocol. Patients were enrolled through a query of the electronic health record (EHR). Single ventricle or functionally single ventricular lesions were excluded. Additionally, only patients with documented blood pressure, height, weight, body mass index (BMI), and echocardiographic data for left ventricular mass (LVM) calculation at the time of their most recent echocardiogram were included. Patients enrolled were classified based on their initial congenital heart disease diagnosis as a newborn (supplemental Figure S-1). The CHD lesions were grouped as tetralogy of Fallot (TOF), transposition of the great arteries (TGA), truncus arteriosus (TA), small left-to-right shunt lesions, congenital pulmonary stenosis, bicuspid aortic valves without stenosis, left ventricular outflow tract (LVOT) obstructive lesions (defined as a primary or residual LVOT Vmax >2 m/s), and minor CHD.

### Echocardiogram Parameters and LV Remodeling

For the purposes of this study, the echocardiographic measurements were measured from M-Mode images, a standard for LV quantification in our echo lab during this timeframe.

Measurements of the interventricular septum (IVSd) and left ventricular posterior wall (LVPWd) were taken at end-diastole. Left ventricular dimensions were taken at end-diastole (LVEDD) and end-systole (LVESD). LVM was determined via the linear method using the formula: 0.8x (1.04x[(IVS+LVID+PWT)^3^-LVID^3^] + 0.6 grams. ^27^ LVM was then indexed to body height to the power of 2.7 (LVM-Ht^2.7^) to adjust for differences in body size ^28–30^. LV function was assessed by left ventricular shortening fraction (LVSF). ^27, 31^

### Blood Pressure Definitions and Categories

Patients were stratified by BP category using the most current American Heart Association/American College of Cardiology guidelines. ^32^ Categories included normotensive (NT, systolic blood pressure (SBP) <120 mmHg), elevated blood pressure (E-BP, 120≤SBP<130 mmHg), stage 1 HTN (HTN-1, 130≤SBP<140 mmHg), and stage 2 HTN (HTN-2, SBP≥140 mmHg), and these measurements were obtained utilizing the blood pressure taken at the time of their most recent echocardiogram. Manual BP were obtained by cardiology clinic nurses trained in acquiring blood pressures. The prevalence of LVH was reported in each group using cutoffs of LVM-Ht^2.7^ ≥ 51 g/Ht^2.7^ given association with cardiovascular disease outcomes with this method^29^ and current guideline cutoffs of >115 g/m^2^ for males and >95 g/m^2^ for females (LVMi). ^27^

### BMI stratification

Patients were categorized by BMI based on the classification from the Centers for Disease Control (CDC) prevention categories, such that underweight: BMI <18.5, healthy weight: BMI 18.5≤ - <25, overweight: BMI 25≤ - 30, and obese: (BMI ≥30).

### Statistical analysis

Demographic, anthropometric, clinical, and echocardiographic parameters were summarized as count (percent) for categorical features and median (Q1, Q3) for continuous features.

Wilcoxon rank sum test was utilized for pre-specified comparisons of echocardiographic parameters between NT and each of E-BP, HTN-1, and HTN-2 patients. P-values were adjusted for multiple comparisons using Holm’s correction method and significant differences in values of echocardiographic parameters was determined at α= 0.05.

Correlation between LVM indexing methods were assessed overall and within levels of BMI category by Pearson’s correlation coefficient and were presented with 95% Confidence intervals (CI). Trends in LVM-Ht^2.7^ for changing values of LVMi were visualized using trend lines generated by univariable linear regression models.

Univariable and multivariable linear regression models were utilized to elucidate the association of SBP and BMI on LVMi and LVM-Ht^2.7^. Multivariable models were adjusted for confounders relevant to the specific exposure based on clinical expertise and analysis of directed acyclic graphs (DAGs) (supplemental Figure S-2). For models with SBP as the primary exposure, adjusted confounders included age, sex, cardiac diagnosis, and BMI. For models with BMI as the primary exposure, adjusted confounders included age, sex, and cardiac diagnosis.

Model estimates for primary exposures of interest were interpreted as mean increase or decrease in LVMi/LVM-Ht^2.7^ for a ten-unit change in the exposure and were presented alongside 95% CI and p-values. Appropriateness of model fit was examined via residual and QQ plots. Additional sets of linear models were implemented to determine which combination of factors were able to explain the greatest amount of patient variation in LVH, as measured by adjusted R2. Values of R2 were interpreted as the proportion of variation in LVH (ranging from 0%-100%) explained by the model, after accounting for the number of included covariates. All statistical analyses were performed in R version 4.2.2 (R Core Team Vienna, Austria).

## Results

A total of 1152 subjects met the inclusion criteria with a mean (SD) age of 24 years (Q1-Q3, 20-32 years) and an equal percentage of males and females. The majority of the cohort was white (88%) (Table 1). BMI and body surface area (BSA) trended higher in the abnormal blood pressure groups. The most common CHD types in this studied population included small left-to-right lesions (27%), and left ventricular outlet obstructions (25%), followed by tetralogy of Fallot (18%), congenital pulmonary stenosis (11%), bicuspid aortic valve disease without stenosis (6.8%), transposition of the great arteries (3.3%), and truncus arteriosus (1.1%).

**Table 1:** Demographics.

| Characteristic | Overall,<br>N = 1,152* | Normotensive,<br>N = 519* | Elevated,<br>N = 313* | Stage I HTN,<br>N = 183* | Stage II<br>HTN,<br>N = 137* |
| --- | --- | --- | --- | --- | --- |
| **Age (years)** | 24 (20, 32) | 24 (20, 31) | 24 (20, 30) | 25 (20, 33) | 26 (20, 39) |
| **Sex** |  |  |  |  |  |
| Male | 577 (50%) | 202 (39%) | 180 (58%) | 115 (63%) | 80 (58%) |
| Female | 575 (50%) | 317 (61%) | 133 (42%) | 68 (37%) | 57 (42%) |
| **Race** |  |  |  |  |  |
| White | 1,014 (88%) | 447 (87%) | 278 (89%) | 167 (91%) | 122 (90%) |
| Black or African American | 108 (9.4%) | 55 (11%) | 26 (8.3%) | 14 (7.7%) | 13 (9.6%) |
| Hispanic | 20 (1.7%) | 9 (1.7%) | 9 (2.9%) | 2 (1.1%) | 0 (0%) |
| Other | 4 (0.3%) | 4 (0.8%) | 0 (0%) | 0 (0%) | 0 (0%) |
| Unknown | 6 | 4 | 0 | 0 | 2 |
| **Height (cm)** | 167 (160, 176) | 163 (156, 171) | 170 (163, 176) | 172 (164, 179) | 169 (164, 178) |
| **Weight (kg)** | 73 (61, 87) | 67 (57, 79) | 74 (63, 86) | 81 (70, 98) | 89 (75, 104) |
| **Body Surface Area (m<sup>2</sup>)** | 1.86 (1.67, 2.06) | 1.76 (1.60, 1.93) | 1.87 (1.72, 2.04) | 1.97 (1.84, 2.20) | 2.08 (1.89, 2.28) |
| **Body Mass Index (kg/m<sup>2</sup>)<sup>†</sup>** | 26 (22, 31) | 24 (21, 30) | 25 (22, 30) | 28 (24, 33) | 31 (25, 36) |
| **BMI Category** |  |  |  |  |  |
| Underweight | 64 (5.6%) | 39 (7.5%) | 18 (5.8%) | 5 (2.7%) | 2 (1.5%) |
| Healthy Weight | 455 (39%) | 240 (46%) | 133 (42%) | 51 (28%) | 31 (23%) |
| Overweight | 278 (24%) | 115 (22%) | 81 (26%) | 51 (28%) | 31 (23%) |
| Obese | 355 (31%) | 125 (24%) | 81 (26%) | 76 (42%) | 73 (53%) |
| **Systolic Blood Pressure (mmHg)** | 121 (112, 130) | 111 (105, 116) | 124 (122, 126) | 133 (131, 136) | 147 (142, 153) |
| **Diastolic Blood Pressure (mmHg)** | 68 (61, 75) | 64 (59, 70) | 70 (64, 76) | 73 (66, 79) | 76 (68, 86) |
| **Diagnosis<sup>‡</sup>** |  |  |  |  |  |
| TOF | 205 (18%) | 101 (19%) | 50 (16%) | 32 (17%) | 22 (16%) |
| TGA | 38 (3.3%) | 15 (2.9%) | 13 (4.2%) | 6 (3.3%) | 4 (2.9%) |
| Truncus | 13 (1.1%) | 4 (0.8%) | 5 (1.6%) | 3 (1.6%) | 1 (0.7%) |
| Small Left to Right Shunts | 311 (27%) | 156 (30%) | 79 (25%) | 48 (26%) | 28 (20%) |
| PS | 128 (11%) | 56 (11%) | 42 (13%) | 13 (7.1%) | 17 (12%) |
| LVOT/Arch Obstruction | 285 (25%) | 95 (18%) | 80 (26%) | 62 (34%) | 48 (35%) |
| Minor CHD | 94 (8.2%) | 52 (10%) | 22 (7.0%) | 10 (5.5%) | 10 (7.3%) |
| BAV without Stenosis | 78 (6.8%) | 40 (7.7%) | 22 (7.0%) | 9 (4.9%) | 7 (5.1%) |

\*Median (Q1, Q3); n (%)
<sup>†</sup>BMI categories based on the classification from the Centers for Disease Control (CDC)
Prevention categories, such that underweight: BMI <18.5, healthy weight: BMI 18.5 ≤ - <25, overweight: BMI 25 ≤ - 30, and obese: (BMI ≥30).
<sup>‡</sup>CHD diagnoses: TOF = tetralogy of Fallot, TGA = transposition of the great arteries, Truncus = truncus arteriosus
PS = pulmonary stenosis, BAV = bicuspid aortic valve

By BP, 45% of patients were NT, 27% E-BP, 16% HTN-1, and 12% HTN-2. Patients in HTN-1 and HTN-2 groups had larger LVPWd and IVSd measurements compared to NT subjects (Table 2). Those in the E-BP, HTN-1, and HTN-2 groups had higher LVEDD values compared to NT. LVM as an absolute value, as well as when as indexed by both BSA and Ht^2.7^, were significantly elevated in only the HTN-2 groups compared to NT patients. Systolic function as quantified by LVSF was preserved and consistent across blood pressure categories.

**Table 2:** Echocardiographic measurements by blood pressure category.

| Characteristic | Overall,<br>N = 1,152* | Normotensive,<br>N = 519* | Elevated,<br>N = 313* | Stage I HTN,<br>N = 183* | Stage II HTN,<br>N = 137* | Significance |
| --- | --- | --- | --- | --- | --- | --- |
| **LVEDD (cm)** | 4.90 (4.50, 5.40) | 4.80 (4.40, 5.30) | **5.00 (4.60, 5.40)** | **5.17 (4.78, 5.50)** | **5.03 (4.80, 5.50)** | *E, I, II* |
| **LVESD (cm)** | 3.11 (2.80, 3.50) | 3.05 (2.70, 3.40) | 3.20 (2.80, 3.54) | 3.20 (2.90, 3.55) | 3.20 (2.80, 3.50) |  |
| **LVPWd (cm)** | 0.81 (0.70, 0.92) | 0.80 (0.70, 0.90) | 0.80 (0.70, 0.90) | **0.90 (0.80, 1.00)** | **0.90 (0.80, 1.00)** | *I, II* |
| **IVSd (cm)** | 0.90 (0.80, 1.00) | 0.84 (0.73, 1.00) | 0.90 (0.78, 1.00) | **0.90 (0.80, 1.00)** | **0.90 (0.80, 1.10)** | *I, II* |
| **LVM (g)** | 145 (115, 184) | 132 (106, 169) | **145 (115, 184)** | **169 (131, 200)** | **173 (140, 212)** | *E, I, II* |
| **LVMi (g/m<sup>2</sup>)** | 78 (65, 94) | 76 (63, 92) | 77 (62, 94) | 83 (68, 97) | **84 (70, 100)** | *II* |
| **LVMi-Ht<sup>2.7</sup> (g/ht<sup>2.7</sup>)** | 36 (29, 45) | 36 (28, 43) | 34 (28, 43) | 37 (32, 47) | **41 (34, 51)** | *II* |
| **LVSF (%)** | 36.2 (32.7, 40.4) | 36.0 (32.4, 40.1) | 35.4 (32.5, 39.5) | 37.0 (33.2, 40.7) | 37.8 (33.3, 41.7) |  |
| **RWT** | 0.35 (0.30, 0.40) | 0.35 (0.30, 0.39) | 0.34 (0.30, 0.39) | 0.34 (0.31, 0.39) | **0.36 (0.32, 0.42)** | *II* |

\*Median (Q1, Q3)
E = Holm-corrected p-value < 0.05 for normotensive vs. elevated
I = Holm-corrected p-value < 0.05 for normotensive vs. stage I hypertension
II = Holm-corrected p-value < 0.05 for normotensive vs. stage II hypertension
LVEDD = left ventricle end diastolic dimension, LVESD = left ventricle end systolic dimension,
IVDSd = interventricular septal systolic dimension, LVM = left ventricle, LVMi = left ventricle
mass indexed to BSA, LVM-Ht<sup>2.7</sup> = left ventricle mass index to allometric height to the power of
2.7, LVSF = left ventricle shortening fraction, RWT = relative wall thickness

A high proportion of LVH by both indexing methods was seen in the HTN-2 group (Table 3). The relative frequencies of hypertrophy and remodeling by LVMi and relative wall thickness (RWT) within each BP group demonstrated a trend of increased concentric hypertrophy in the HTN-2 group (supplemental Figure S-3). Notably, a high concordance was demonstrated between the LVM indexing methods (BSA and Ht^2.7^) by BMI category (Figure 1).

**Figure 1:**
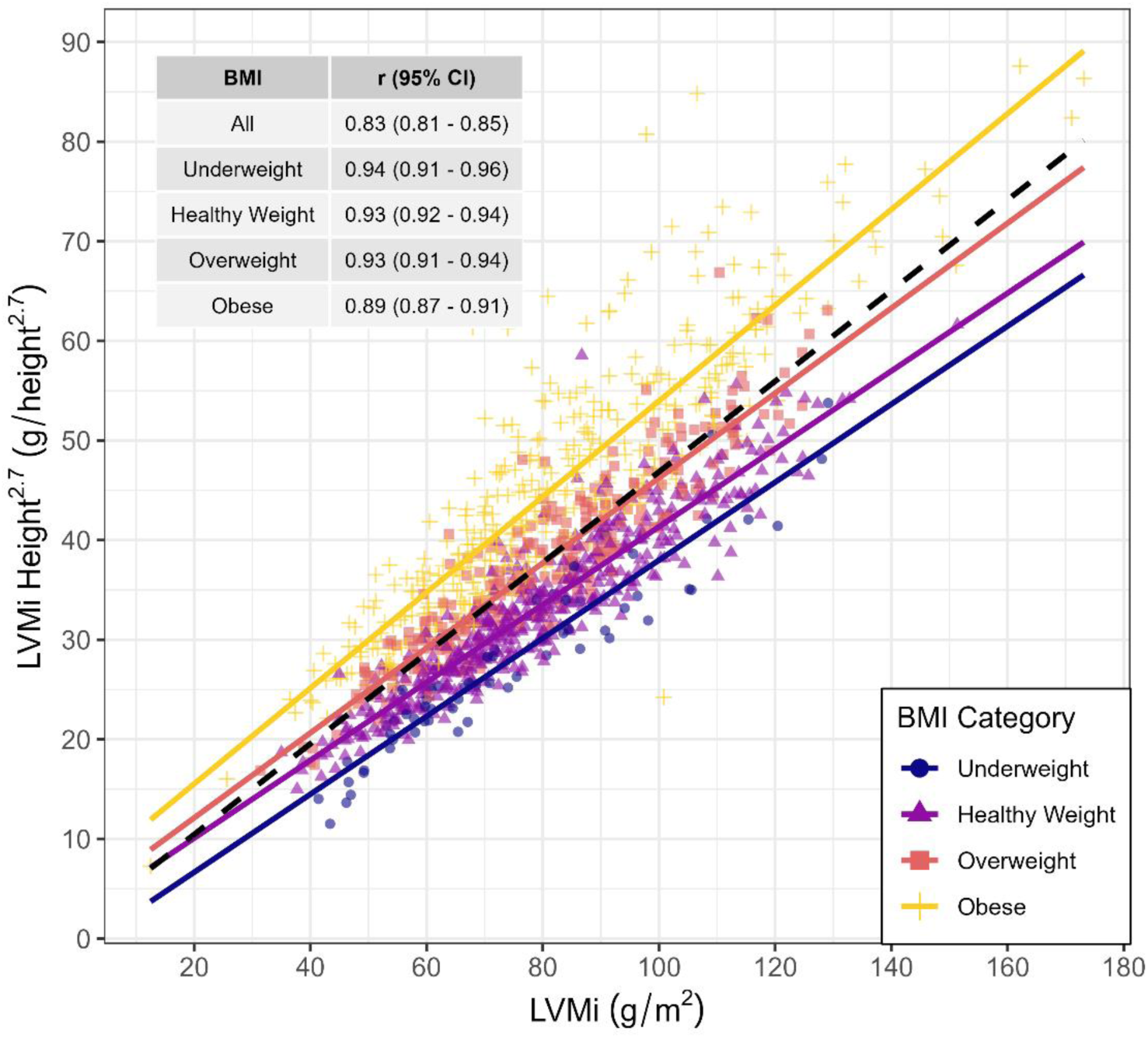
Correlation between LVM indexed to BSA (LVMi) and Height^2.7^ (LVM-ht^2.7^)

**Table 3:**
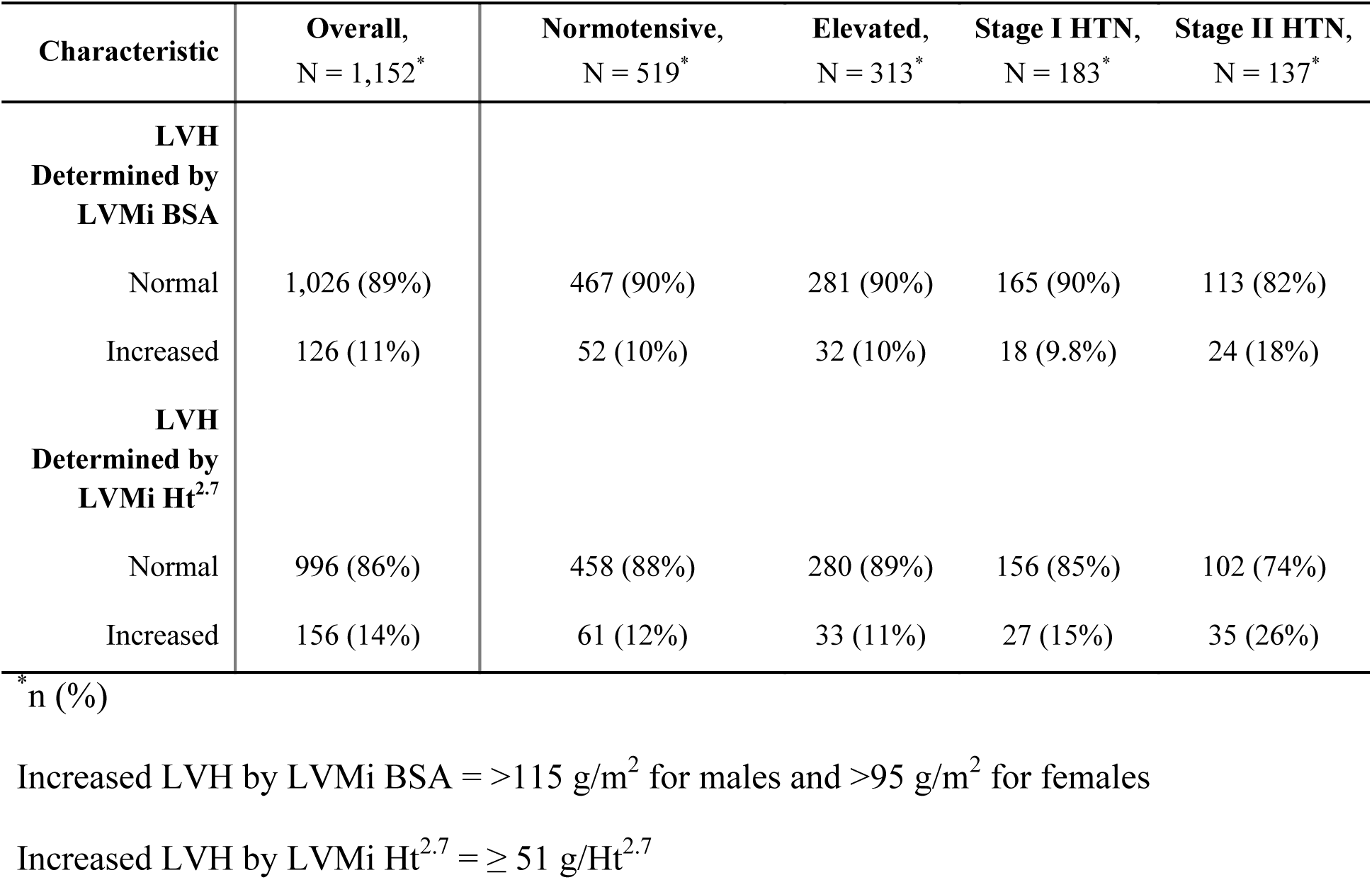
Prevalence of LVH by LVM indexed to BSA (LVMi) and Height^2.7^ (LVM-ht^2.7^)

Univariable and multivariable linear regression models for the impact of SBP and BMI on LVH can be seen in Figure 2, and demonstrates that for every increase of 10 mmHg of systolic blood pressure, there was an associated 2.15 g/m^2^ (95% CI: 1.31, 2.99; p < 0.001) increase in LVMi in the unadjusted model. With the adjusted model, there was a 0.56 g/m^2^ (95% CI: −0.27, 1.4; p = 0.19) increase in LVMi. There was an estimated increase of LVM-Ht^2.7^ by 1.27 g/m^2^ (95% CI: 0.81, 1.73l p < 0.001) in the unadjusted model but −0.15 g/m^2^ (95% CI: −0.56, 0.26; p = 0.48) in the adjusted model. When evaluating BMI, a 10-unit increase of BMI was associated with an increase in LVMi by 1.93 g/m^2^ (95% CI: 0.2, 3.66; p = 0.029) unadjusted and 1.27 g/m^2^ (95% CI: −0.38, 2.93; p = 0.13) when adjusted. For BMI and indexing via LVM-Ht^2.7^, a 10-unit increase of BMI was associated with an LVM-Ht^2.7^ increase of 8.37 g/m^2^ (95% CI: 7.55, 9.18; p < 0.001) unadjusted and 7.95 g/m^2^ (95% CI: 7.13, 8.77; p < 0.001) when adjusted.

**Figure 2:**
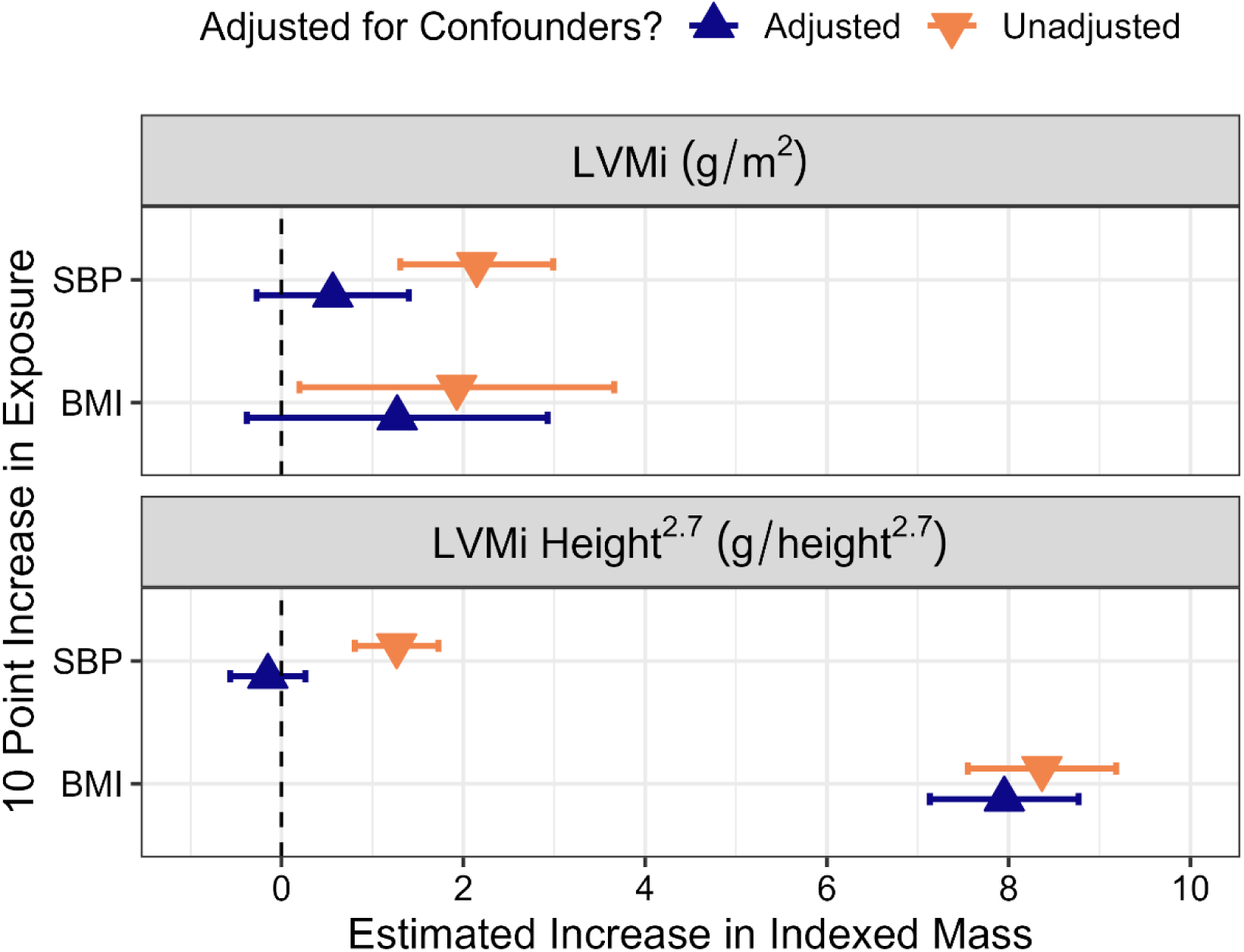
Univariable and Multivariable Linear Regression Models for Indexed LVM. SBP = systolic blood pressure. BMI = body mass index. For models with SBP as the primary exposure, adjusted confounders included age, sex, cardiac diagnosis, and BMI. For models with BMI as the primary exposure, adjusted confounders included age, sex, and cardiac diagnosis.

In addition to the primary models with covariates chosen by examination of DAGs, sets of linear models with covariates iteratively added were run to elucidate which factor(s) most explained variation in LVH (Figure 3). All model specifications generally showed low explained variation in both LVM indexed to BSA and LVM-Ht^2.7^ by examination of R^2^ values. Regardless of combinations of relevant model parameters, only 12%-15% of variation in LVMi could be explained. Explained variation in LVM-Ht2.7 ranged from 6%-8% for models with baseline features included and either known LVOTO or SBP. However, the explained variation increased substantially, reaching 29%, when an individual’s BMI was known. Inclusion of all relevant covariates only increased explained variation in LVM-Ht2.7 by 1% as opposed to knowledge of just baseline characteristics and BMI.

**Figure 3:**
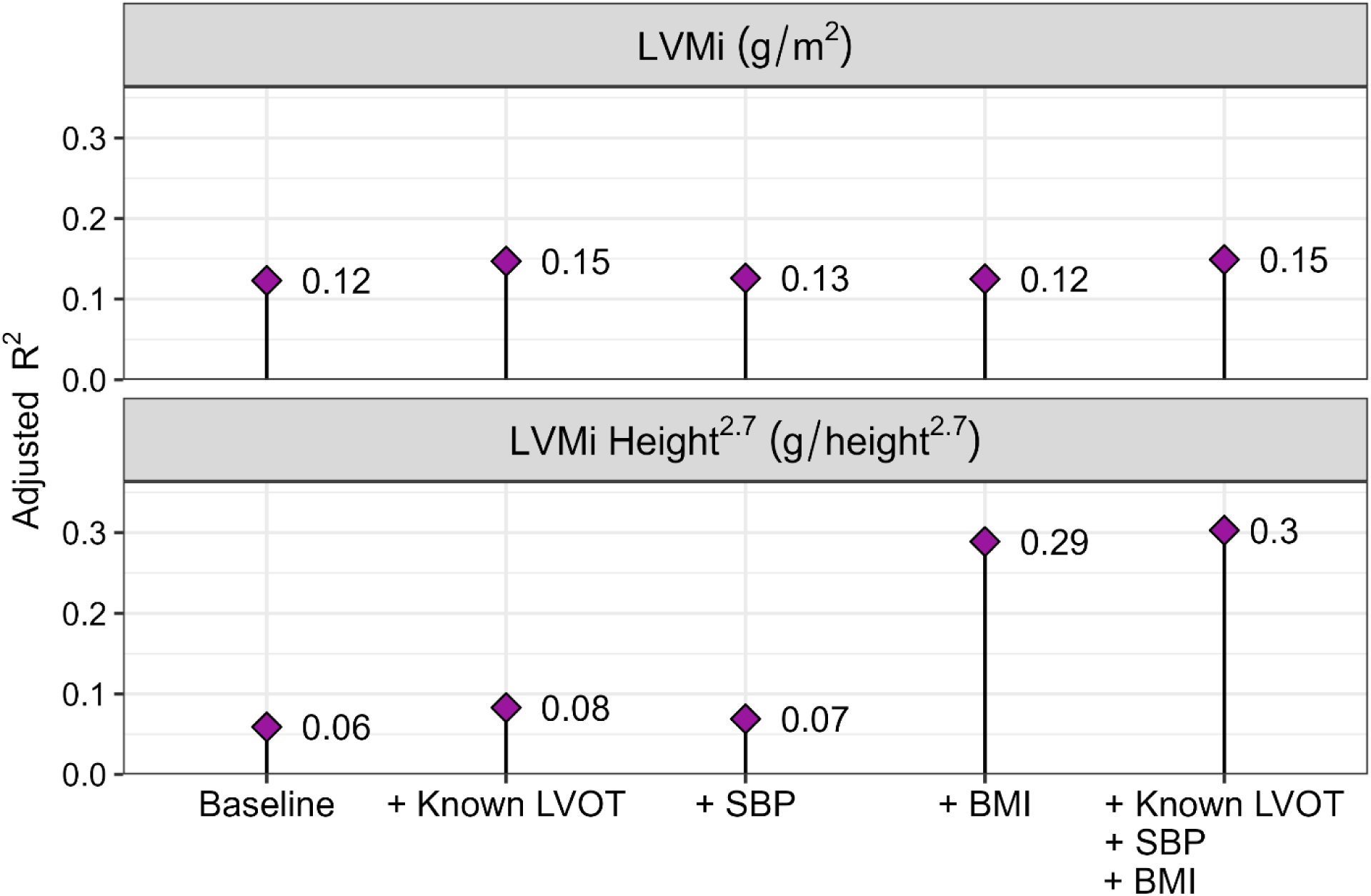
Adjusted R^2^ values from linear models demonstrating low-explained variation in both LVMi and LVM-Ht^2.7^ for combinations of relevant clinical factors. SBP = systolic blood pressure. BMI = body mass index. Baseline models included factors of: age and sex.

## Discussion

This study included a large population of adults with a wide variety of CHD anomalies, and provides a unique retrospective analysis on correlates of obesity and BP on the development of LVH. Findings from this study highlight that obesity has the greatest impact on the development of LVH in ACHD subjects.

We found that in this large cohort of ACHD patients, 55% had abnormal blood pressures, higher than reported for the general population, ^33^ which may reflect the possible impact of the underlying CHD on vascular tone. ^34^ High blood pressure was a risk factor for increased LVH, especially in HTN-2 groups. LVM indexed to both BSA and LVM-Ht^2.7^ appeared to be higher in those with higher BP, similar to adults and youth without CHD. ^35, 36^ As HTN in adults without CHD has historically been known to predict adverse cardiovascular events such as stroke, myocardial infarction and systolic and diastolic dysfunction, ^32, 37–39^ it has also been associated with development of LVH, which is itself a predictor of cardiovascular events. ^17, 40–42^ By correlation methods in this study, elevated BP was less impactful for the development of increased LVM, however. Regarding CHD lesion type, those with small left-to-right shunt lesions and LVOT or aortic arch obstruction CHD had higher prevalence of higher blood pressures, consistent with prior data that focused on left sided obstructive lesions. ^24, 43–45^

Just as obesity has been historically known as a significant independent predictor of LVM in adults without CHD, ^46^ weight continues to prove itself to be an important factor in the development of LVH in this study. Notably, BMI was the single most impactful variable in predicting LVM, and was found to significantly increase LVH when indexing LVM to Ht^2.7^, even after adjusting for confounding factors. While the same was not found when assessing LVM indexing to BSA, this discrepancy may be a factor of weight being included in BSA formula. Regardless, prior research showing LVH identified by LVM normalized for height to allometric power has been shown to adequately predict CV events. ^29, 30^ Interestingly, patients grouped as having an LVOT obstruction did not explain a significant amount of one’s variation in LVH as compared to BMI. This may reflect improving surgical advances over many generations of CHD operations, such as high success rates of neonatal coarctation of the aorta repairs in the neonatal population.^47^ In addition, there’s improving hypertension management such as the use of ambulatory blood pressure monitoring, as well as evolving interventional catheterization procedural methods including stenting. At 30% maximally explained variation, there is still a large amount of variability in this patient population regarding LVH.

Obesity has a high prevalence, including within the ACHD population. Similar to this study, recent literature highlights that BMI percentile was associated with LVH in pediatric and adolescent patients.^23^ Another study has reported the prevalence of overweight/obesity in the ACHD population being 65%, ^48^ underscoring the important of having measurable factors such as LVH may prove beneficial for clinical care for the ACHD population, given current data supporting that regression in LVM leads to lesser risk of CV disease in adults without CHD. ^49^ Future studies focusing on the long-term implications and outcomes of LVM in the ACHD population are paramount in this regard.

This study broadens our understanding of the impact of obesity and HTN in the ACHD population, and highlights the need for early detection of obesity to prevent progression of LVH. Considering the obesity epidemic in the general population worldwide and within the ACHD population, ^8, 48, 50^ the impact of obesity on LVH development is underscored in this study. As ACHD patients require long term follow up from a young age, there are ample opportunities to intervene on both elevated blood pressures and weight management.

## Limitations

While our study includes a large sample of patients, as a single center retrospective cohort study, determination of causation of LVH is limited. The ejection fraction was measured for many patients, but not consistently reported to be included in this analysis, so LVSF was used instead as a marker for left ventricular systolic function. We believe our large sample size, inclusive of all types of biventricular CHD, mitigated selection bias, although we did not analyze the types of CHD in a multivariate format. Given this, we are unable to comment on specific CHD variations which may carry a higher impact on LVM. Given the descriptive nature of our work, whether patients were diagnosed and were being actively treated for HTN was not investigated. Additionally, single time-point blood pressures were utilized for our analysis. Serial blood pressure evaluation and incorporation of ambulatory blood pressure monitoring data would further enhance our understanding of the true prevalence of HTN in this cohort and provide insight into the timing of development of end-organ damage. Future studies should be aimed at longitudinal changes in weight and blood pressure and LVH and the impact of pharmacological intervention and social determinants on LVM, as well as outcomes.

## Abbreviations

ACHD: Adult Congenital Heart Disease
CHD: Congenital Heart Disease
LVMI-ht2.7: Left ventricular mass indexed to patient height to the power of 2.7
LVMi: Left ventricular mass indexed to patient body surface area
NT: Normotensive
E-BP: Elevated blood pressure
HTN-1: Stage 1 hypertension
HTN-2: Stage 2 hypertension
SBP: systolic blood pressure
LVOT: Left ventricular outflow tract
LVSF: Left ventricular shortening fraction
TOF: Tetralogy of Fallot
TA: Truncus arteriosus

## Data Availability

All data referred to in this manuscript is accessible upon request.

## Acknowledgements

None

## Supplemental Index

**Figure S1:**
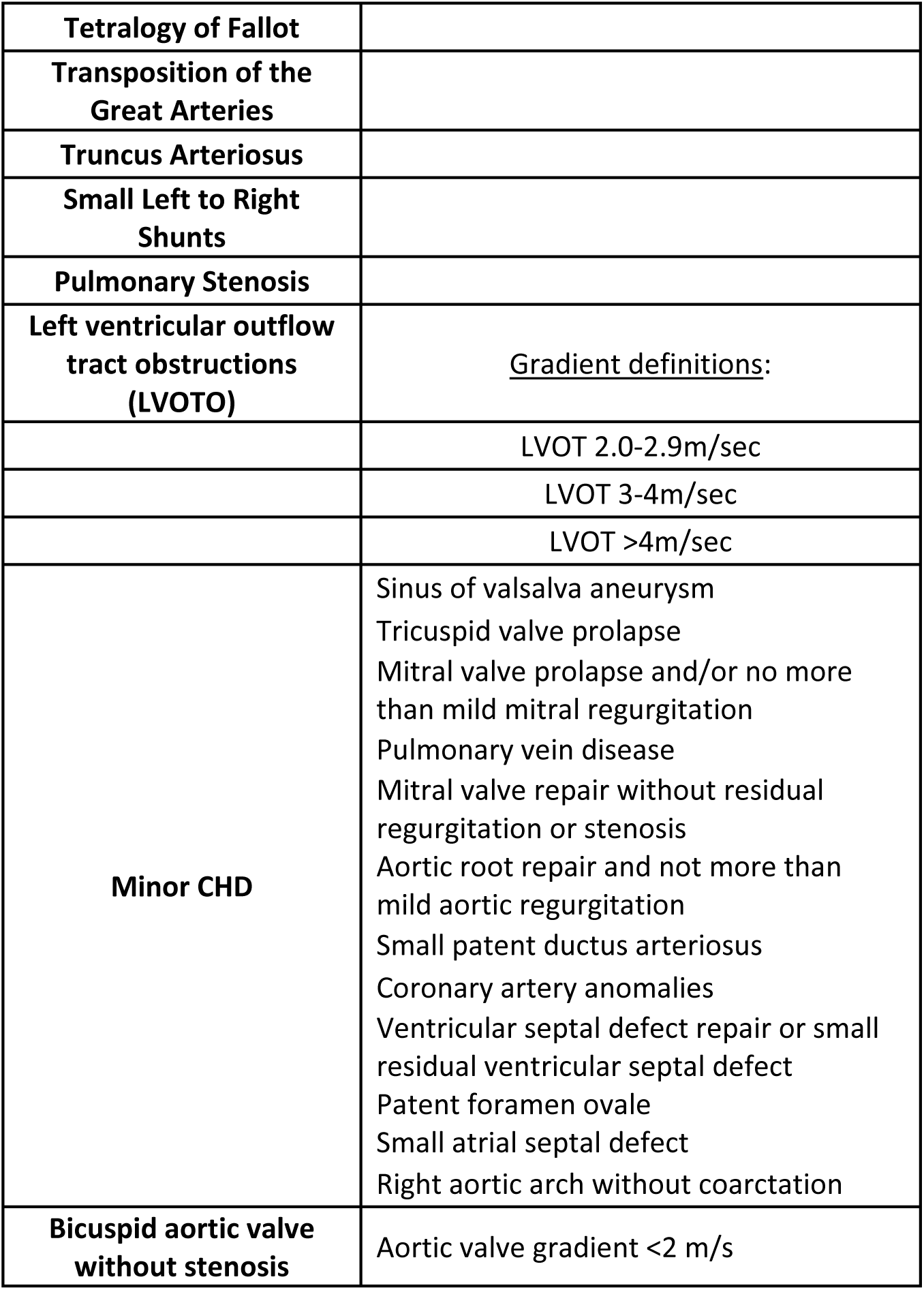
Categorization of CHD

**Figure S2:**
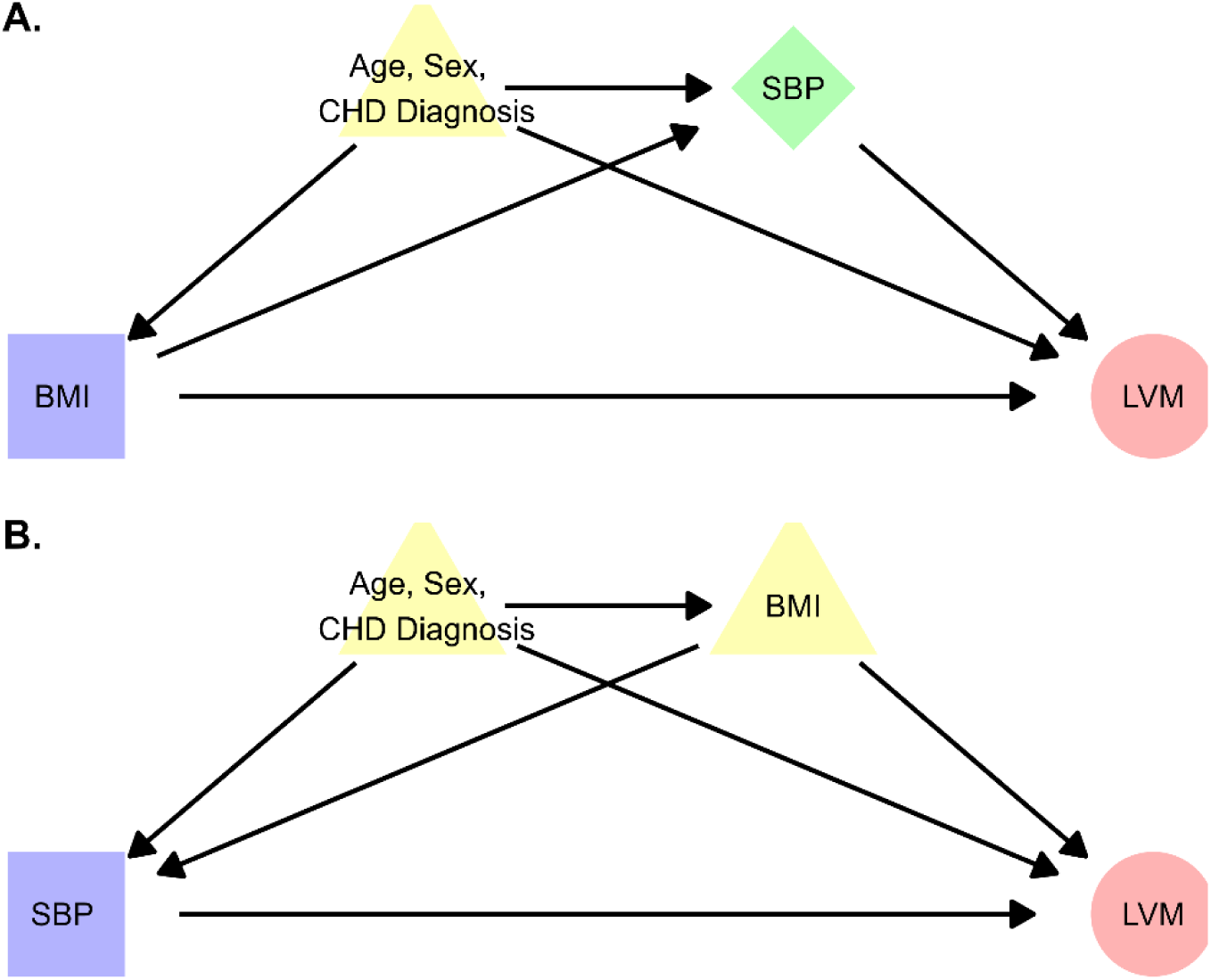
Directed acyclic graphs (DAGs) for the effect of BMI and systolic blood pressure on LVM.

**Figure S3:**
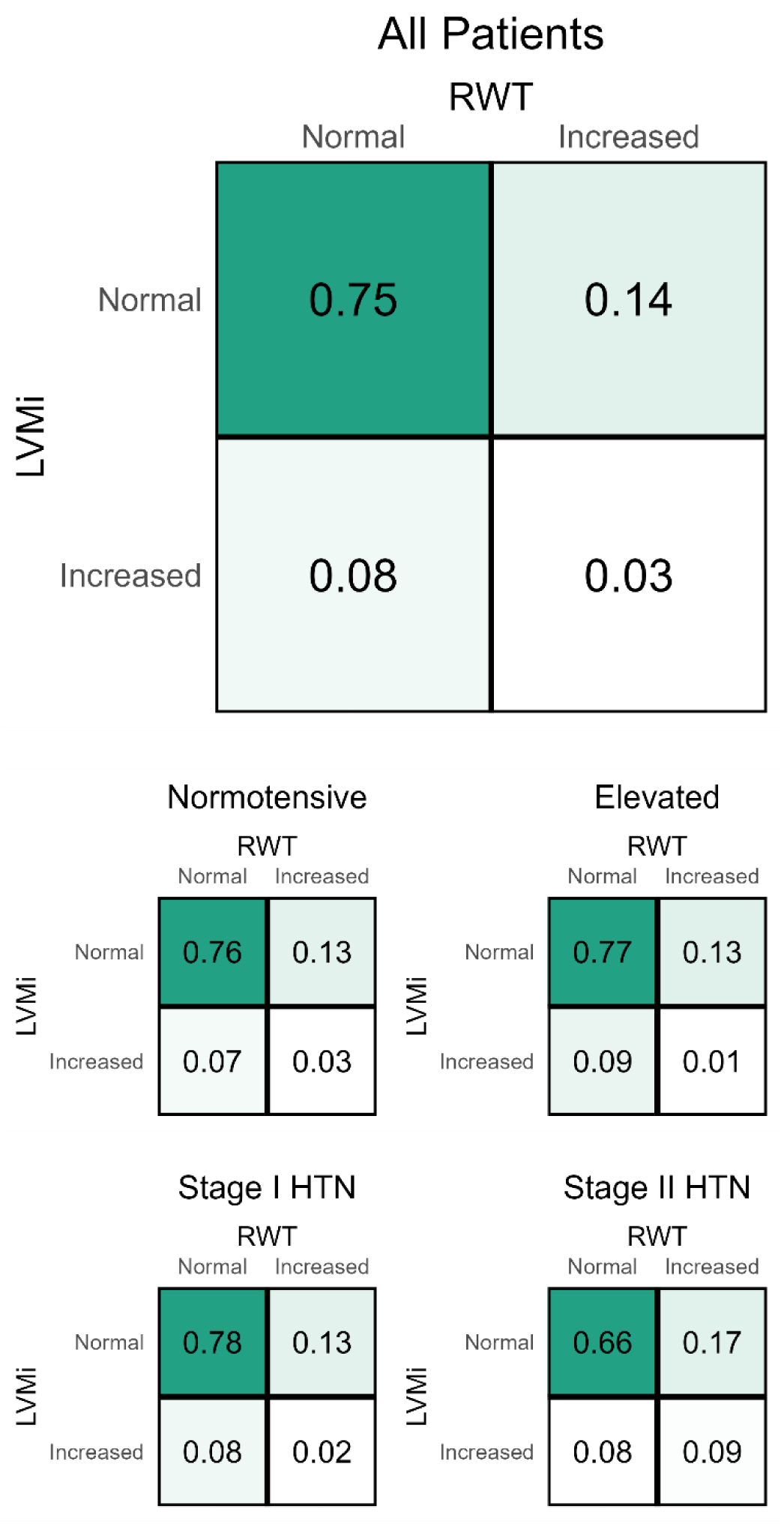
The relative frequencies of hypertrophy and remodeling by LVMi and relative wall thickness (RWT) within each BP group.

